# Designing an Electronic Patient Reported Outcomes Information Infrastructure Supported by the RE-AIM Implementation Framework

**DOI:** 10.1101/2025.03.31.25324980

**Authors:** Amy C. Moreno, Angela Peek, Toshiko S. Stein, Kevin R. Shook, Sara M. Ali, Laia Humbert-Vidan, Aileen Chen, Miriam Lango, Anna Lee, Michael Spiotto, William H. Morrison, Adam S. Garden, Jack Phan, Steven J. Frank, Katherine A. Hutcheson, David I. Rosenthal, Clifton D. Fuller, G. Brandon Gunn

## Abstract

**Objective:** Patient-reported outcomes (PROs) contain valuable information that can be leveraged by providers to perform timely interventions and improve quality of life and survival. However, the implementation of electronic PROs (ePROs) remains a challenge from technical, behavioral, and evaluation perspectives. Our objective was to construct a robust electronic health record (EHR)-integrated ePRO information infrastructure founded on RE-AIM (Reach-Effectiveness-Adoption-Implementation-Maintenance) principles.

**Materials and Methods:** We used Epic Systems as our EHR platform to build the MD Anderson Symptom Index-Head and Neck Module (MDASI-HN) for release to all patients undergoing evaluation and/or treatment in our HN Radiation Oncology clinics. RE-AIM metrics were established and used to design patient-, provider-, and implementing facilitator information tools.

**Results:** From January 2021 to July 2024, our ePRO program has collected 13,156 *patient-submitted* ePROs on 3,497 unique HN patients, with a 12-month sustained ePRO compliance rate of 82%. We also propose a dynamic clinical implementation-evaluation cycle. This model can be used to continuously (re)define, build, and adapt ePRO information tools for patients, providers, and program facilitators.

**Discussion:** Our ePRO framework has several benefits including integrated clinical data for enhanced decision-making, potential scalability, and use of a common EHR system. Formative (i.e., mid-phase) evaluation and feedback were essential in our program, allowing for timely optimization of ePRO compliance, ePRO usage by clinical staff, and secondary use of high-quality ePRO data.

**Conclusion:** In this article, we provide a valuable roadmap towards developing a comprehensive, EHR-based ePRO information infrastructure simultaneously optimized for clinical utility and implementation evaluation founded on RE-AIM principles.

## INTRODUCTION

### Background and Significance

The value of patient-reported outcomes (PROs) in oncology has been well established in healthcare literature. PROs are formally defined as the status of patient’s health condition that comes directly from the patient.^1^ PROs can characterize a spectrum of multidimensional domains including overall quality of life (QOL), functional status (e.g., swallowing function post-radiotherapy [RT] for head and neck cancer), symptom burden, and patient experience.^2^ When collected prospectively and longitudinally throughout a cancer survivors’ lifetime, dynamic patterns of early and late PRO burden can be investigated to identify high-risk populations and develop more effective, data-driven toxicity mitigation strategies. ^3–7^ Clinically, PROs have also been associated with enhanced multidisciplinary care, patient QOL, symptom management, and patient-provider communication.^8–10^

Despite the abundance of support for adopting PROs, high PRO compliance can be challenging to achieve even for heavily resourced, multicenter cancer clinical trials. Independent predictors of PRO completion include institutional size, patient age, and treatment group.^11^ Additional barriers to PRO implementation in clinical practice include issues with organizational workflows, limited stakeholder engagement (i.e., clinical staff and patients), time management, and information technology infrastructures that are underdeveloped for PRO collection, visualization, and efficient clinical reporting.^12–14^

Electronic health record (EHR) systems present a unique opportunity to construct and implement *integrated, electronic* PROs (ePROs) with structured metadata (i.e., who, when, and how were ePROs captured). Moreover, relevant clinical data can be embedded within the same ePRO information system (i.e., imaging, laboratory, and treatment data) to provide additional context to reported symptoms. Given rising interests in EHR-based ePRO integration, the Patient Centered Outcomes Research Institute (PCORI) developed guidelines for healthcare providers interested in ePRO adoption;^15^ however, these recommendations are generalized to enable versatile applications in different settings, subsequently leaving knowledge gaps on how to monitor and ensure the maintenance, or institutionalization, of ePROs.

Several implementation evaluation frameworks, such as RE-AIM, have been well established in public health assessments of community-based interventions and are being increasingly applied in cancer settings.^16–21^ The RE-AIM framework, for example, conceptualizes the impact of implementation efforts as a product of five domains. These include reach (proportion of target population participation and risk characteristics), effectiveness (positive and negative outcomes of an intervention and variability across subgroups), adoption (representativeness settings and personnel that will adopt the interventions), implementation (fidelity to and/or adaptations to the intended interventions), and maintenance (extent to which interventions are sustained over time or institutionalized).^22,23^

Reports on RE-AIM-based ePRO implementation efforts or the technical build of EHR-based ePRO systems exist.^24–27^ However, these tend to be smaller scale studies or discrete publications without discussions of information tools that can be generated to support the various stages of an implementation program. To address this knowledge gap, we systematically outline our work in designing an EHR (Epic)-based ePRO information infrastructure. We also describe our implementation evaluation cycle that supports an ongoing large cancer center ePRO implementation program driven by the RE-AIM framework.

## MATERIALS AND METHODS

### The Electronic PRO Implementation Program

An Epic-based ePRO implementation program using the RE-AIM evaluation framework was initiated on January 28, 2021 at the MD Anderson Cancer Center Department of Radiation Oncology, Head and Neck Section. The target cancer population to receive ePROs were all patients with a diagnosis of head and neck cancer (HNC). The target setting was outpatient visits in the Head and Neck Radiation Oncology (HNRO) clinic. The selected ePRO was the MD Anderson Symptom Index-Head and Neck Module (MDASI-HN), a 28-item validated questionnaire designed to measure general and HNC-specific symptom burden.^28^ The MDASI-HN is rated on a numerical scale ranging from 0 (not present) to 10 (as bad as you can imagine) and has a 24-hour recall period. Additional stakeholders for this implementation study included all clinical staff involved in HNRO patient care, including schedulers (PSCs), medical assistants (MA), nurses, advanced practice providers, and faculty. ACM served as the site facilitator and worked closely with the Epic Analytics team (AP, TS, KS, SA) to develop the ePRO-driven information tools.

From January 28, 2021 to July 31, 2024, 13,156 <u>patient-submitted</u> ePROs have been collected on 3,497 unique HNC patients. A critical all-staff meeting occurred in February 2023 that formally launched the RE-AIM information infrastructure. As a result, ePRO compliance accelerated from 35% to 87.4% with sustained monthly ePRO completion average of 82% over the last 12 months. The RE-AIM framework and proposed metrics for this study are described below. This study had Institutional Review Board approval under protocol 2024-0002.

### The RE-AIM Framework

An overview of the framework and interactive RE-AIM planning tool can be found at re-aim.org. The target population for this initiative (i.e., **Reach**) has been described above. The ePRO Program was approved by the institution’s Patient Survey Informatics Committee (PSIC) who provides oversight of ePRO builds and proposed workflows. The program was then promoted internally through the dissemination of clinical staff educational materials (Supplement S1), quarterly ePRO meetings, and patient-facing educational materials (Supplement S2). **Effectiveness** was defined as total ePRO compliance which can be calculated as a percentage of the completed ePROs compared to the total of assigned ePROs. Patient adherence to completing longitudinal ePROs throughout their cancer journey from consultation to on-treatment weekly “see” visits (WSVs) and/or follow ups was also examined. Key characteristics to review the degree of ePRO **Adoption** included the clinical setting (i.e., the main center or the proton therapy center [PTC]) and individual provider clinics (i.e., the assigned physician and their staff). The feasibility of ePRO **Implementation** was expected to be high as EHR-integrated ePROs can be automatically assigned to eligible visit orders (i.e., a future follow up) and be automatically delivered to patients via Epic MyChart® in preparation for their future visit(s). Several staff-facing information tools were also designed to facilitate staff-specific responsibilities in the designed ePRO clinical workflow. Proposed adjustments or modifications to the ePRO program were recorded during monthly-to-quarterly meetings with the Analytics Team and ePRO Committee, the latter inclusive of all HNRO clinical staff representatives. Changes to the EHR-based criteria could be audited by examining changes to the rule-based logic. **Maintenance** of EHR-based ePRO integration was routinely monitored through automated reports developed using the Web Intelligence (WebI) tool which is part of the SAP Business Intelligence Platform and retrieves data from the Epic Clarity database.^29,30^

## RESULTS

### EHR-based Information Architecture Design

Figure 1 illustrates the information infrastructure designed to support EHR-based ePRO implementation into clinical practice. This clinical implementation-implementation evaluation cycle facilitates the optimization of the initiative by automating RE-AIM planning procedures and global metrics.

**Figure 1.**
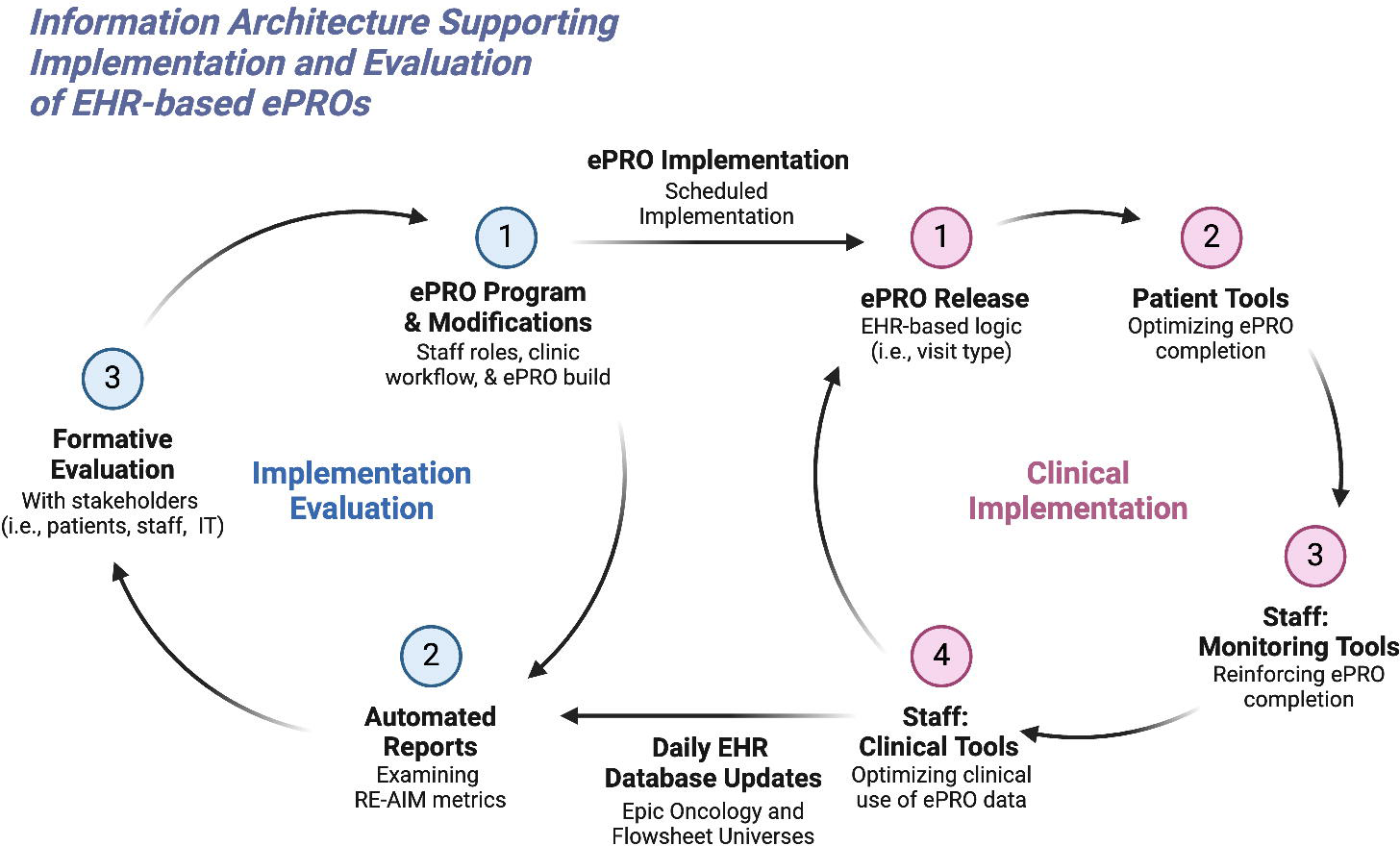
Information Architecture for EHR-based ePRO Integration.

### ePRO Build and Release

Upon defining the target population, setting, and ePRO tool (i.e., MDASI-HN), we collaborated with EHR analysts to build structured flowsheets. We subsequently created patient-entered questions from individual flowsheet rows and grouped items into a new questionnaire. Each question was reviewed by the facilitator for the potential designation of high alert values (HAVs) which are defined as patient-entered scores above a certain threshold that should trigger a rule-based action linked to a Best Practice Advisory (BPA). For example, if a patient rated the question “your shortness of breath at its worst” as 8 or greater, the proposed BPA was to route these HAVs to a nursing staff pool in-basket for prompt triaging. All nurses were educated on how to manage incoming HAVs pre-implementation. A total of 5 items in the MDASI-HN were assigned as HAV-containing questions (Q1, Q5, Q6, Q16, Q19. In addition, high symptom interference scores were routed to a designated social worker.

A combination of various rules was developed to assign EHR-based ePROs to patients. For simplicity, ePROs were linked to a series of approved visit types including new patient, consult, WSV, and follow up. Telehealth visit codes were also included as this implementation study was initiated during the COVID-19 pandemic. The deployment timeframe was based on the validated questionnaire’s recall period (i.e., 24 hours).

### Patient-facing Tools for ePRO Completion

Activated ePROs can be completed by patients through MyChart®, Epic’s proprietary patient web portal and mobile application.^31^ MyChart® enables automated notifications to patients of upcoming visits with their clinical providers. When logged into the patient portal, patients can access visit-linked ePROs by 1) clicking on the *Menu* button and then *Questionnaires* or 2) clicking on their scheduled visit and finding the section titled “Prepare for Your Visit” (Figure 2).

**Figure 2.**
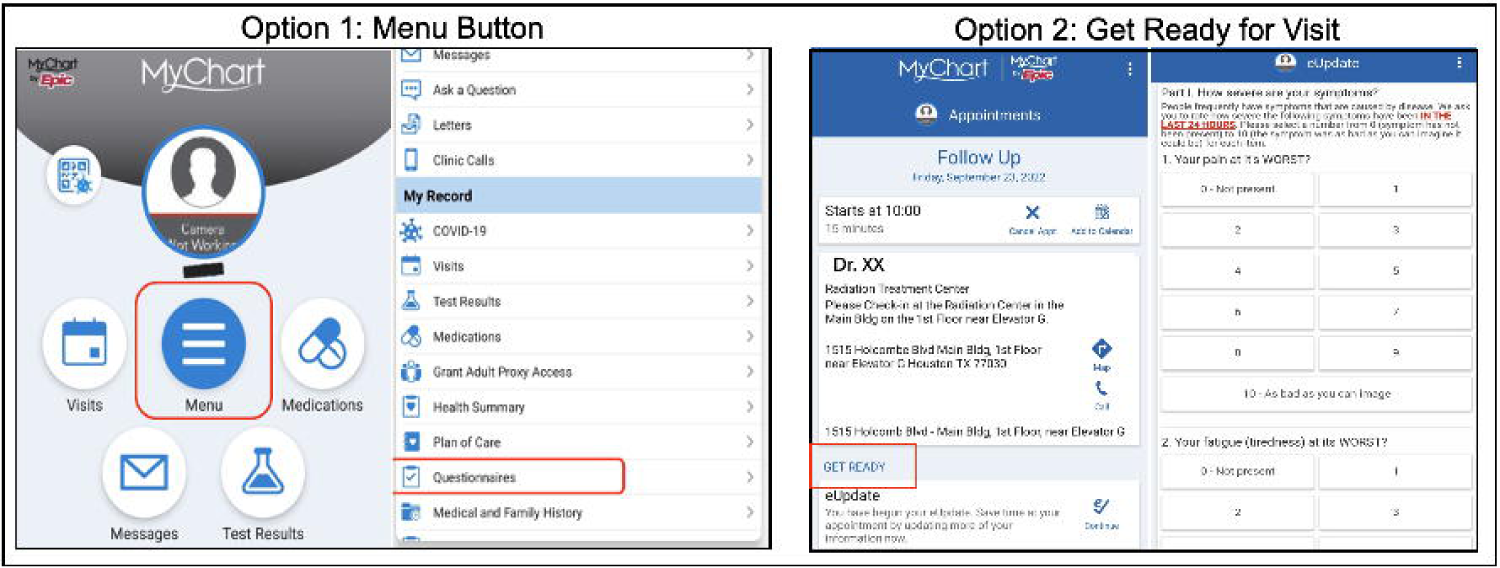
EHR-based ePRO Patient Access Points.

Epic’s Welcome application is a valuable tool for patient-entered ePROs at the time of check-in. This application can be installed on iPads, android tablets, or kiosks placed near the registration desk, and enables patients to complete assigned or ad-hoc ePROs. Schedulers with formal training and Epic access can also assign new or add-on ePROs real-time (i.e., for different visit types or criteria) by generating a QR code in Epic. This allows staff to link the ePRO to the visit and can be completed on a Welcome tablet to subsequently flow data into the established ePRO flowsheet in Epic. In our program, Welcome-enabled iPads have played a key role in the scalability and maintenance of our ePRO initiative.

### Clinical Staff Tools for Monitoring and Reinforcing ePRO Completion

Continuous, real-time monitoring of ePRO completion status by clinical stakeholders is critical for improving and maintaining an ePRO implementation program. Clinic schedule properties are customizable in Epic as shown in Figure 3. Users can right click on their schedule to review its properties, search for the ePRO status column (labeled per facilitator/Epic team preference) in the *Available Columns* section, add it to their schedule (green box), and move its position using the up and down arrows (blue box) in the *Selected Columns* section. Figure 3B shows the *MDASI-HN RO* ePRO status column located on the staff’s schedule with various values available including “*Assigned”* (not started)*, “Started at Home”* (partially completed), and “*Completed.”* This schedule column was integrated into all staff schedules to facilitate daily screening of ePRO compliance. Schedulers checking patients into clinic were then given the role to remind patients with an *Assigned* status to complete ePROs on their mobile phone or the Welcome tablet prior to being roomed. Of note, patients with a status of “*Completed, Completed”* had 2 ePROs assigned and completed for that visit (i.e., MDASI-HN and the OHIP questionnaires, the latter of which evaluates oral health function).^32^

**Figure 3.**
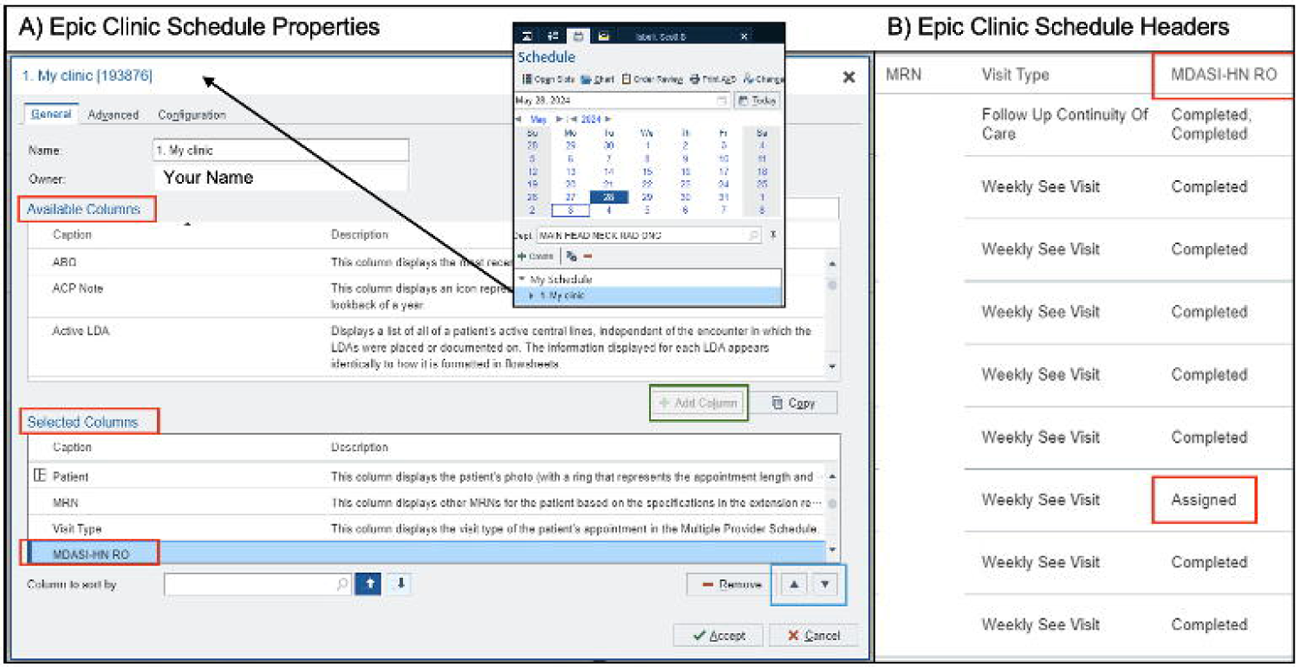
Schedule Properties and ePRO Status Column.

For patients having issues with technology or lacking a smart mobile device, and clinics without the Welcome application, designated clinical staff can assist with entering ePROs manually through the flowsheet tab in the EHR hyperspace (Figure 4). The flowsheet tab within an open patient encounter can display a variety of flowsheets, including the activated ePRO labeled here as *Symptom Inventory MDASI-HN*. Patient-entered data (i.e., via MyChart® or the Welcome application) will showcase a human icon to the left of the value. Questions missed or incomplete PROs can be quickly identified and reviewed with the patient for manual completion by clinical staff as highlighted in Figure 4 where there is a dash to the left of the newly entered value.

**Figure 4.**
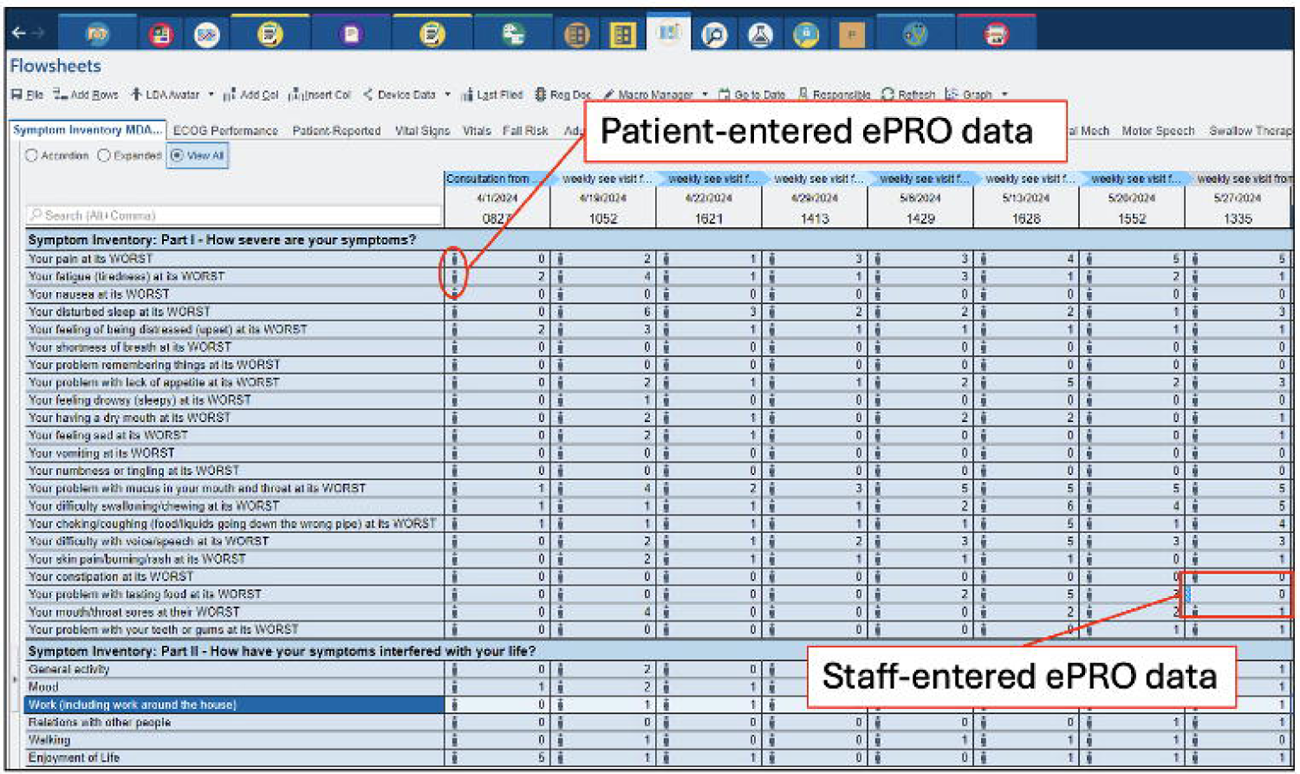
EHR Integrated ePRO Flowsheets.

### Clinical Staff Tools for ePRO Integration in Clinical Decisions and Documentation

EHR-based ePROs provide versatile options for analyzing and using ePRO data during clinical assessments, symptom management, and documentation (Figure 5). Automated ePRO in-basket messages are valuable for timely and prioritized patient triaging as they are immediately deployed to designated staff (i.e., nursing pool) if and when HAVs are entered by a patient. Figure 5A shows an in-basket message displaying the time of ePRO completion. The highlighted values in black are linked to HAV-enabled questions, and the highlighted red values with an exclamation point denote when a HAV threshold has been met. SlicerDicer (Figure 5B), a data exploration tool in Epic, can be leveraged by clinical staff to review general statistics on ePRO collection using selected parameters (i.e., the number of HAV-containing responses based on visit type). The customizable, interactive Synopsis display is perhaps one of the most powerful tools for providers as it can integrate temporal ePRO data with other clinical data. Per feedback from our HNRO clinical team, the Synopsis display (Figure 5C) was designed to visualize vital signs first followed by ePRO data, laboratory data (i.e., CBC and CMP), systemic therapy (i.e., agent, dose, and delivery dates), and imaging data (i.e., CT and MRI scan reports). This enriched ePRO context allows for quicker pattern recognition and more data-driven proactive symptom management (i.e., recommending anti-nausea medications if vitals and nausea scores significantly increase after chemotherapy delivery in specific patients). Customizable trending is an additional function within the Synopsis tab. For example, vitals and specific ePRO items can be trended together as shown in Figure 5C where a patient’s weight is displayed alongside the reported pain score during RT and afterward during routine cancer surveillance visits. Trended items have a graph to the right of their name (yellow circles) and HAVs can still be quickly identified in this display (red box).

**Figure 5.**
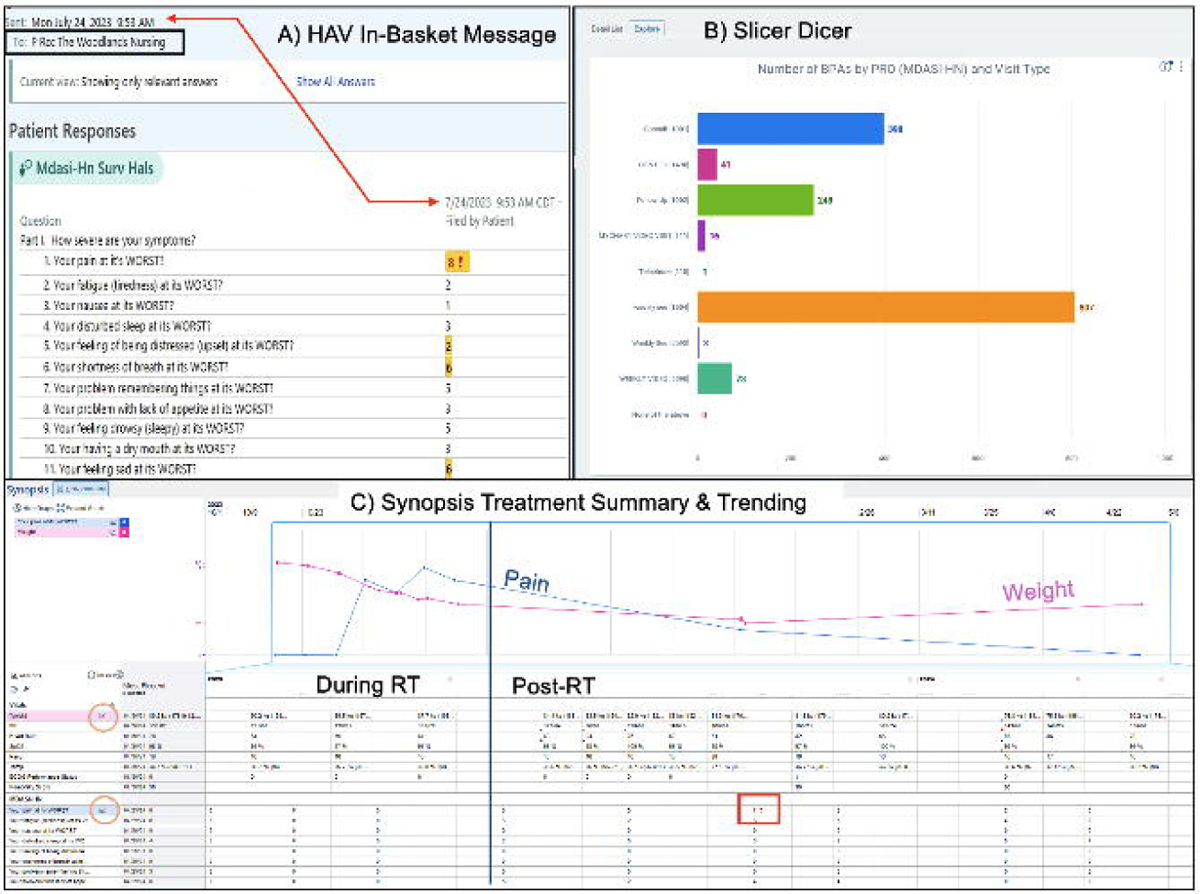
Clinical Tools for Analyzing ePROs.

As ePRO data is linked to a unique flowsheet identifier, several additional tools can be developed from EHR-integrated ePROs to expedite clinical documentation needs and ordering. Firstly, flowsheet-linked dot phrases, or SmartText, can be preset into a note template to automatically import a series of completed ePROs into clinical notes with aggregate score values included if applicable (Supplemental Figure 1). Secondly, remote (i.e., between visits) symptom monitoring can be readily implemented by developing order-based, not visit-based, questionnaires. These orders can be incorporated into a larger SmartSet or Express Lane of orders with frequency of remote ePRO release pre-defined based on specific conditions. For example, our remote ePRO series order, which is nestled under a larger ‘Follow Up Order SmartSet’, is built to remotely deploy weekly MDASI-HN questionnaires for two months post-RT to track and address symptom burden peak not otherwise captured in current standard of care practice.

### Designing Customized Reports for Implementation Evaluation

Pre- and post-implementation needs assessments are necessary to improve the effectiveness of customized, automated reports. Implementation facilitators can utilize these assessments to provide constructive feedback to their teams. Feedback content and frequency of communication depends on the targeted stakeholder and stage of the implementation, respectively. For example, facilitators may want to share ePRO compliance metrics and demographic data of responders versus non-responders with all clinical staff (i.e., stakeholders) more frequently at the launch of an ePRO program and then transition to general ePRO compliance rates on a quarterly basis during the maintenance phase after addressing major barriers. In our study, we were able to develop multiple reports in WebI detailing the *Reach* of the program by linking flowsheet data to patient demographics, the *Effectiveness* by tracking ePRO compliance, the *Adoption* by filtering reports by location and providers, and the *Implementation-Maintenance* phase by tracking the sustainability of ePRO compliance over longer periods of time. Figure 6 shows temporal ePRO completion rates, including an initial observational period coinciding with the COVID-19 pandemic.

**Figure 6.**
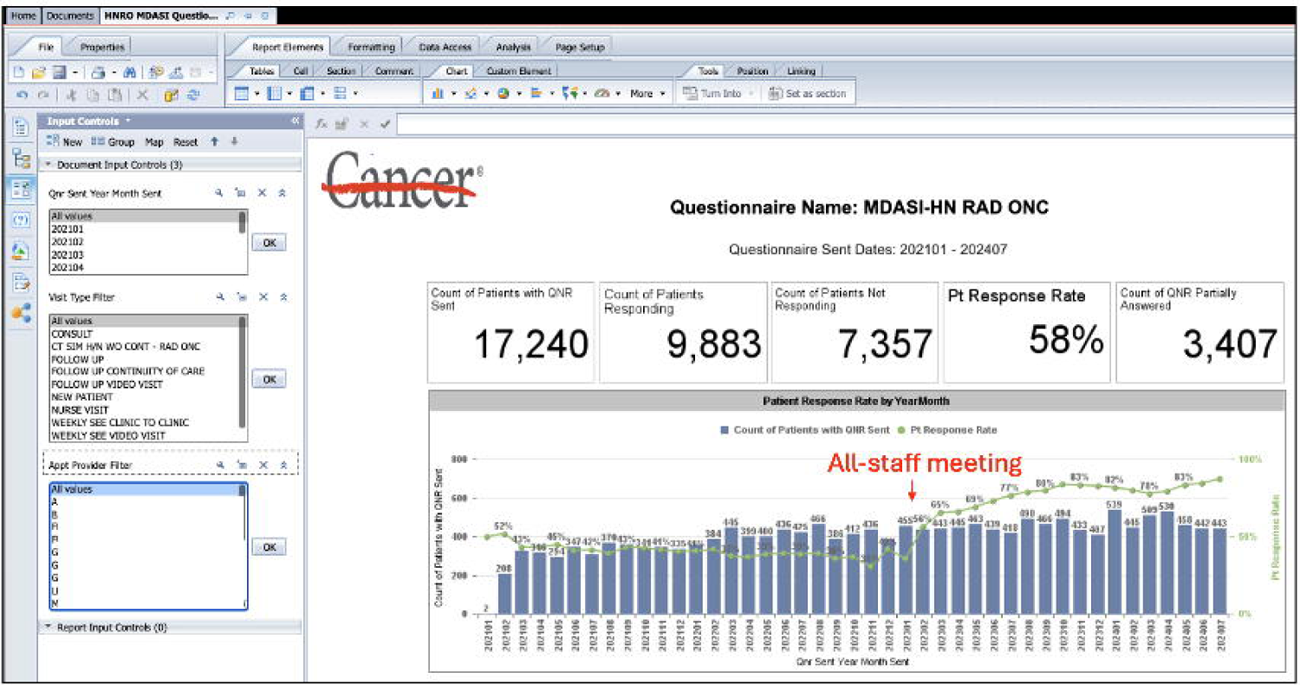
WebI Report on ePRO Implementation Reach.

When the Welcome tablets were launched later in the program in March 2024 (main center) and June 2024 (PTC), there was a need to evaluate the utilization of the tablets and whether the rate of ePRO completion was improving over time. An EHR-based workbench report was generated for more frequent reporting of such data. This demonstrated on average about 22% ePRO compliance for several months using this method of ePRO delivery (Supplemental Figure 2).

### Performing Routine Formative Evaluation

While most implementation studies provide an end-of-program summative evaluation with feedback on whether the results met the stated goals of the program,^33^ formative evaluation enables mid-course modifications or adaptations to the program to improve the intervention. For *implementation-focused* evaluations, the HNRO ePRO Committee met quarterly or more frequently to discuss direct observations and identify barriers to implementation. For example, these meetings provided insight on new visit codes that had been added to Epic but had not been included in the ePRO logic which resulted in staff schedules showing several visits without a linked ePRO. This information was used to develop a two-phased resolution plan: 1) notify PSCs to check for visits with missing ePROs, log and report the associated visit codes, and assign QR codes on tablets for ePRO completion at time of patient check-in, and 2) coordinate a timely update of the EHR criteria logic to include new visit codes. For *progress-focused* evaluations, the WebI ePRO completion report was automated for dissemination to facilitators (ACM) on a monthly basis. The facilitator then used the information as positive reinforcement for clinics with high patient engagement and as a time resource reallocation tool to focus on identifying different barriers affecting clinics with low ePRO compliance.

## DISCUSSION

There is a significant need to disseminate methodologies on how to build, maintain, and utilize comprehensive healthcare information architectures that support all phases of ePRO implementation programs. To our knowledge, this is the first report to summarize such an infrastructure developed using RE-AIM principles and one of the top 5 most adopted EHR systems in the world. Over 325 million patients have electronic health records in Epic Systems (Epic Systems Corporation),^34^ making an Epic-based ePRO system strategic for enabling scalability, translatability across disciplines, and usability of ePROs in clinical decision-making processes via an ‘all-in-one’ clinical data access environment.

When designing an ePRO implementation program, the **ePRO delivery mechanism** should be carefully considered as it impacts resource allocation and scalability of the program. For example, a large academic radiation oncology department performed a 2-year ePRO feasibility study using Tonic as an ePRO software with demonstrated quick ePRO data transfer to the EHR within two minutes after completion.^35^ However, ePRO delivery was highly dependent on tablet dissemination (18 iPads distributed throughout clinics) and personnel oversight as MAs had to select the patient on the tablet, then the disease site and desired ePRO prior to handing the tablet over to the patient. To prevent issues with clinical workflow, MA appointments were often created to accommodate time for ePRO collection. In such circumstances, the scalability and translatability of the ePRO program can be substantially hindered by rising resource costs including more personnel, equipment, and physical spaces that some practices or departments may not be able to afford. Our ePRO infrastructure leveraged Epic’s ability to deploy visit (and order)-based ePROs via MyChart so that ePROs could be completed remotely by the patient at any time point within the ePRO’s defined recall period, even up to time of check-in. Furthermore, scaling up our program to capture ePROs on patients with other cancer diagnoses or patients seen in other clinics would require relatively low-cost efforts from our EHR analysts to incorporate additional codes into the ePRO assignment algorithm. In our study, we observed that the Welcome tablets were helpful in increasing ePRO compliance by about 22%. This required a new workflow for our schedulers but was streamlined by the quick generation of QR codes linked directly to patient charts for a given ePRO survey. Therefore, we recommend that within an EHR-integrated ePRO program, tablet-enabled ePROs should be considered as a complimentary, not primary, source of ePRO delivery as they are often tied to additional resource costs. To improve MyChart-enabled ePRO collection, patient educational resources should be developed or revised to emphasize the importance of ePROs as part of high-quality standard of care.

A major barrier towards effective use of individual-level ePRO data can be the **design of ePRO data visualizations and functions.** A recent systematic review found that visualizations lacking patient summaries, graphical visualizations with interactive options (i.e., ability to trend symptoms), and easily distinguishable abnormal results were identified as barriers to ePRO implementation.^14^ Given the direct ePRO integration hero the EHR, our Synopsis window can efficiently package high yield temporal data with several interactive options allowing for filtering, highlighting, and trending of ePRO data alongside key clinical/treatment data. Moreover, HAVs are easily distinguishable within the Synopsis window and are can be quickly communicated to providers via the automated ePRO notification and rerouting system, thereby facilitating prompt triaging and interventions. Another provider task that is expedited via this ePRO platform is clinical documentation through the imbedding of ePRO-based SmartText phrases within clinical note templates.

In addition to advocating for the use of a formalized evaluation framework to identify specific metrics for an ePRO initiative, we propose a harmonious, two-cycle implementation model of dynamic information needs, optimization, and utilization that is supported by a comprehensive information infrastructure. In the clinical implementation cycle, patient-facing interfaces as well as staff-facing information tools for reinforcing ePRO compliance and ePRO data usage in clinical decision-making processes are prioritized and optimized via continuous feedback from all stakeholders with pre-defined roles in the implementation initiative. Since Epic-based ePRO data is collected in a structured format and deposited daily within the Clarity reporting database,^29^ facilitators of ePRO implementation programs can leverage this near real-time data through connected analytical software (i.e., WebI) that generate automated reports of RE-AIM metrics.^21^ These reports subsequently become the preliminary feedback during formative evaluations to identify new or persistent barriers to ePRO adoption that need to be addressed.

Limitations to our study include the use of only one EHR system which has not been adopted by all institutions or independent providers. Epic also may provide customizations to fit institutional needs, thereby limiting potential ePRO interoperability (e.g., ePRO flowsheets constructed differently). However, several EHR vendors are increasingly providing their own clinical outcomes assessment tools like Oracle’s PRO instruments which are directly integrated with patient clinical data.^36^ Regardless of the EHR system in place, we have presented a comprehensive methodology for developing an EHR-agnostic information infrastructure that facilitates ePRO adoption and implementation evaluation. Lastly, our study did not incorporate computerized adaptive testing (CAT) which aims at reducing items administered and patient-related survey fatigue.^37^ Additional work is required to build this functionality in major EHR software and develop EHR API integrations to allow for more seamless data sharing without the need of constant modifications or real-time error monitoring if either the EHR or the API requires a substantial update.^25^

## CONCLUSION

Our study is synergistic with prior and ongoing ePRO initiatives, and we provide a valuable roadmap towards developing a comprehensive, EHR-based ePRO information infrastructure simultaneously optimized for clinical utility and implementation evaluation founded on RE-AIM principles. Future directions include expansion of this ePRO framework on a multidisciplinary level to standardize the prospective collection of ePROs in various discipline settings with alternating surveillance schedules (e.g., head and neck surgery, medical oncology, oral oncology). Moreover, given the passive and scalable collection of high-quality structured data, our secondary data usage aim will be to develop data-driven toxicity risk prediction models for operationalizing surveillance schedules and formulating algorithmic symptom management pathways during and after multimodal cancer therapy.

---

Supplemental Figure 1. SmartText Example.

Supplemental Figure 2. EHR Reporting Workbench PRO Completion Report.

## Pre-print Availability Statement

In accordance with NIH Policy NOT-OD-17-050, *Reporting Preprints and Other Interim Research Products,* which specifies: “The NIH encourages investigators to use interim research products, such as preprints, to speed the dissemination and enhance the rigor of their work”, we have deposited a pre-peer review version of the initial version of this submitted manuscript at medrxiv with DOI available upon acceptance.

## Data Availability Statement

All data produced in the present work regarding information infrastructure design are contained in the manuscript.

## Ethics Approval Statement

The IRB of The University of Texas MD Anderson Cancer Center approved this research under protocol 2024-0002.

## CRediT statement

In accordance with the Contributor Roles Taxonomy (CRediT, https://credit.niso.org/), the contributing authors have designated responsibilities and individual author attribution. The corresponding author (ACM) assumes responsibility for role assignment, and all contributors have been given the opportunity to review and confirm assigned roles:

*Conceptualization*: ACM, AL, CDF, DIR, JP, KAH, and GBG; *Methodology*: ACM, CDF, DIR, KAH, GBG; *Validation:* all coauthors; *Resources*: all coauthors; *Data curation*: ACM; *Funding acquisition*: ACM and CDF; *Investigation*: ACM; *Project administration*: ACM; *Supervision*: ACM, CDF, DIR, KAH, and GBG; *Writing – original draft*: ACM; *Writing - review & editing*: all coauthors.

## ICJME Author Statement

In accordance with International Committee of Medical Journal Editors (ICJME, https://www.icmje.org/) recommendations, all authors affirm qualification for authorship via the following criteria: *“Substantial contributions to the conception or design of the work; or the acquisition, analysis, or interpretation of data for the work; AND Drafting the work or reviewing it critically for important intellectual content; AND Final approval of the version to be published; AND Agreement to be accountable for all aspects of the work in ensuring that questions related to the accuracy or integrity of any part of the work are appropriately investigated and resolved.”*

## Disclaimer

The content is solely the responsibility of the authors and does not necessarily represent the official view of the National Institutes of Health.

