## Supplementary figures and images for "Designing an Electronic Patient Reported Outcomes Information Infrastructure Supported by the RE-AIM Implementation Framework"

### Supplemental Figure S1

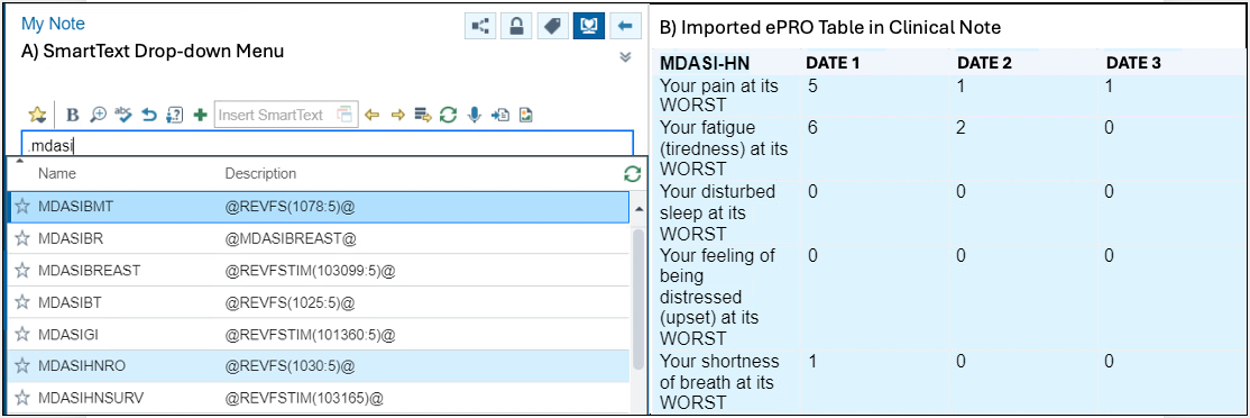

### Supplemental Figure S2

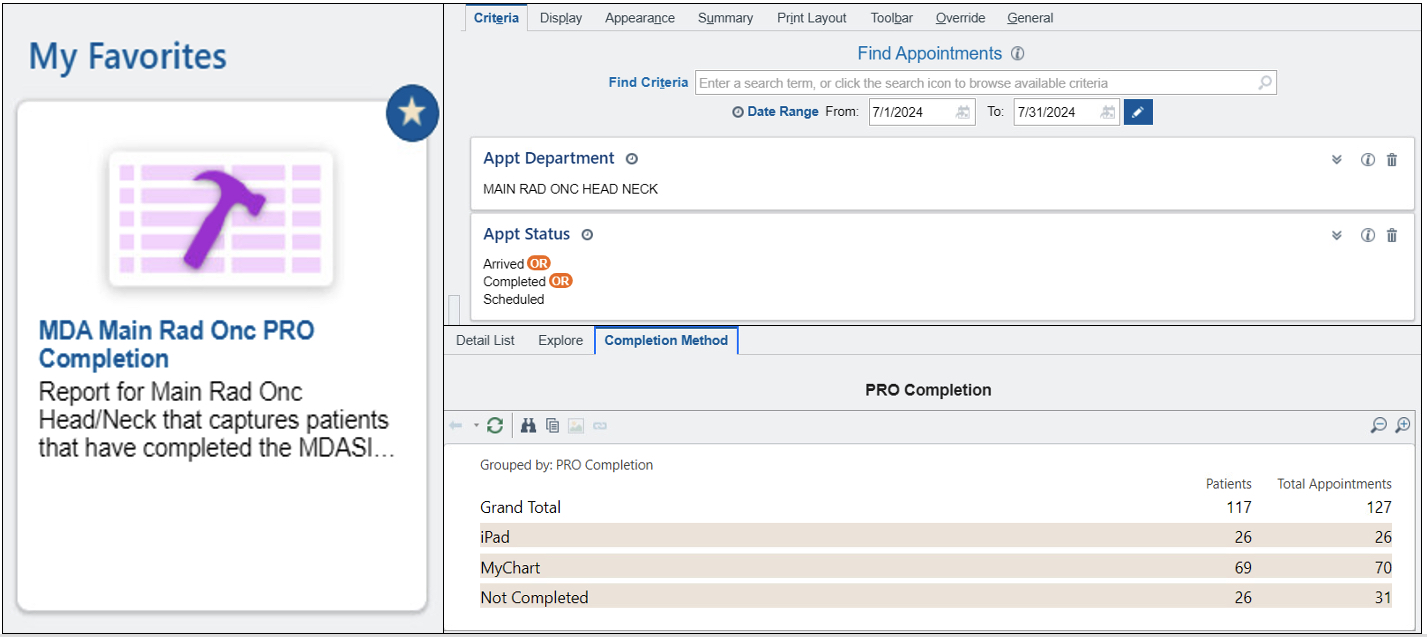
